# Phenotypic variability and Gastrointestinal Manifestations/Interventions for growth in Ogden syndrome (also known as *NAA10*-related Syndrome)

**DOI:** 10.1101/2022.03.16.22272517

**Authors:** Katherine Sandomirsky, Elaine Marchi, Maureen Gavin, Karen Amble, Gholson J. Lyon

## Abstract

Our study of 61 children with Ogden Syndrome, an X-linked disorder due to *NAA10* gene mutations, demonstrated a high prevalence of growth failure, with weight and height percentiles often in the failure-to-thrive diagnostic range; although dramatic weight fluctuations and phenotypic variability is evidenced in the growth parameters of this population. Although never previously explored in depth, the gastrointestinal pathology associated with OS includes feeding difficulties in infancy, dysphagia, GERD/silent reflux, vomiting, constipation, diarrhea, bowel incontinence, and presence of eosinophils on esophageal endoscopy, in order from most to least prevalent. Additionally, the gastrointestinal symptom profile for children with this syndrome has been expanded to include eosinophilic esophagitis, cyclic vomiting syndrome, Mallory Weiss tears, abdominal migraine, esophageal dilation, and subglottic stenosis. Although the exact cause of poor growth in OS probands is unclear and the degree of contribution to this problem by GI symptomatology remains uncertain, an analysis including nine G-tube or GJ-tube fed probands demonstrates that G/GJ-tubes are overall efficacious with respect to improvements in weight gain and caregiving. The choice to insert a gastrostomy or gastrojejunal tube to aid with weight gain is often a challenging decision to make for parents, who may alternatively choose to rely on oral feeding, caloric supplementation, calorie tracking, and feeding therapy. In this case, if OS children are not tracking above the FTT range past 1 year of age despite such efforts, they should promptly undergo G-tube placement to avoid prolonged growth failure. If G-tubes are not immediately inducing weight gain after insertion, recommendations include altering formula, increasing caloric input, or exchanging a G-tube for a GJ-tube by means of a minimally invasive procedure. Future directions could include a prospective natural history study investigating whether G/GJ tube insertion affects the cognitive trajectory, rate of reaching developmental milestones, and GI symptomatology of OS children in a positive or negative manner.

## Introduction

The purpose of our study is to explore the clinical phenotype of Ogden Syndrome with a focus on its gastrointestinal manifestations. Current scientific literature reports the following GI-related complications associated with this syndrome: poor feeding, dysphagia, aspirations, umbilical hernia, diarrhea (Rope et al. 2011), multiple gastrointestinal infections (Wu and Lyon 2018), vomiting requiring hospitalization (Gogoll et al. 2021), anteriorly displaced anus (Afrin et al. 2020), oversized liver (Maini et al. 2021), chronic constipation (Rasmus Ree et al. 2019), eosinophilic gastritis (Gupta et al. 2019), esophageal and gut dysmotility syndrome, esophagitis, fecal incontinence, silent reflux, and gastric reflux (Cheng et al. 2019). Such gastrointestinal symptoms may play a role in the commonly documented growth findings of postnatal growth failure observed in children with Ogden Syndrome; parallel findings of growth retardation were observed in *NAA10* knock-out: yeast (Mullen et al. 1989; Polevoda and Sherman 2003), zebrafish (*Danio rerio*) (R. Ree et al. 2015), and mice (Kweon et al. 2021; Lee et al. 2019). However, it is worth noting that Ogden Syndrome is phenotypically variable in humans (Wu and Lyon 2018), with certain probands experiencing normal weights and stature (Støve et al. 2018). Multiple probands analyzed in the literature required feeding tube placement to treat feeding difficulties and/or FTT (Gogoll et al. 2021; Afrin et al. 2020; Maini et al. 2021; Gupta et al. 2019; Cheng et al. 2019; Saunier et al. 2016; Bader et al. 2020), but the efficacy of tube feeding is only discussed in two publications, albeit very briefly. According to one such study by Rope et al., a male with a c.109T>C p.(Ser37Pro) variant (Proband 60 in our study) was able to build up subcutaneous fat stores, presumably because of G tube feeding “ in infancy (Rope et al. 2011). A separate study refers to a female *de novo* c. 384T>G p.(Phe128Leu) variant whose early feeding difficulties were improved by G-tube placement at 2 months old, as per her parent, who also stated that, at 1.5 years old, this child’s weight was lower than that of age-matched females (Cheng et al. 2019). However, this was the extent of the discussion regarding the direct benefits or detriments of G-tube feeding, and neither of these studies included growth parameters immediately preceding and following tube insertion. This study aims to expand upon the effectiveness of nasogastric, gastric, and gastro-jejunal tubes on the growth trajectories of Ogden Syndrome probands.

## Methodology

Our methods consisted of mining medical records and data from interviews conducted by Dr. Gholson Lyon (in-person or via videoconferencing with the families of probands analyzed). Oral and written consent was obtained for research and publication, with approval of protocol #7659 for the Jervis Clinic by the New York State Psychiatric Institute - Columbia University Department of Psychiatry Institutional Review Board. Excel and Prism software were utilized for creating graphs and tables. SimulConsult was used to determine weight percentiles and standard deviation values of height and weight parameters. In an effort to exclude identifying information, the term “parent “ was used in lieu of “mother “ or “father, “ “sibling “ was used instead of “brother “ or “sister, “ and age ranges, rather than exact ages, were included throughout this text. Of note, certain probands included in our study have already been described in the literature. Refer to Supplementary Table 1 to determine which probands in our study correspond to previously published OS children. Probands 2, 3, 5, 7-9, 12, 17-19, 22, 25, 27-29, 32-35, 37-50, 52, and 54 have not heretofore been published. These individuals will be described in more detail in a pending manuscript related to the phenotypic spectrum of Ogden Syndrome.

**Table 1.** Prevalence of Gastrointestinal Symptoms in Analyzed Probands.

| **Gastrointestinal Symptom** | **Total Probands with Symptom** | **Percentage of Probands with Symptom** |
| --- | --- | --- |
| **Feeding Difficulties in Infancy** | **52** | **90** |
| **Constipation** | **20** | **34** |
| **Diarrhea/Loose Stools** | **14** | **24** |
| **Vomiting** | **26** | **44** |
| **GERD/Silent Reflux** | **32** | **54** |
| **Dysphagia** | **45** | **75** |
| **Esophageal Eosinophils on Endoscopy** | **6** | **10** |
| **Bowel Incontinence** | **7** | **12** |

## Results

The probands investigated in this study possess varying *NAA10* mutations (refer to Supplementary Table 1 for each individual ‘s cDNA mutation and protein variant).

According to the plotted weight percentile trajectories of our study ‘s probands, there exists a great deal of variability in the growth of children with Ogden Syndrome (Figure 1A). However, upon closer analysis

**Figure 1.**
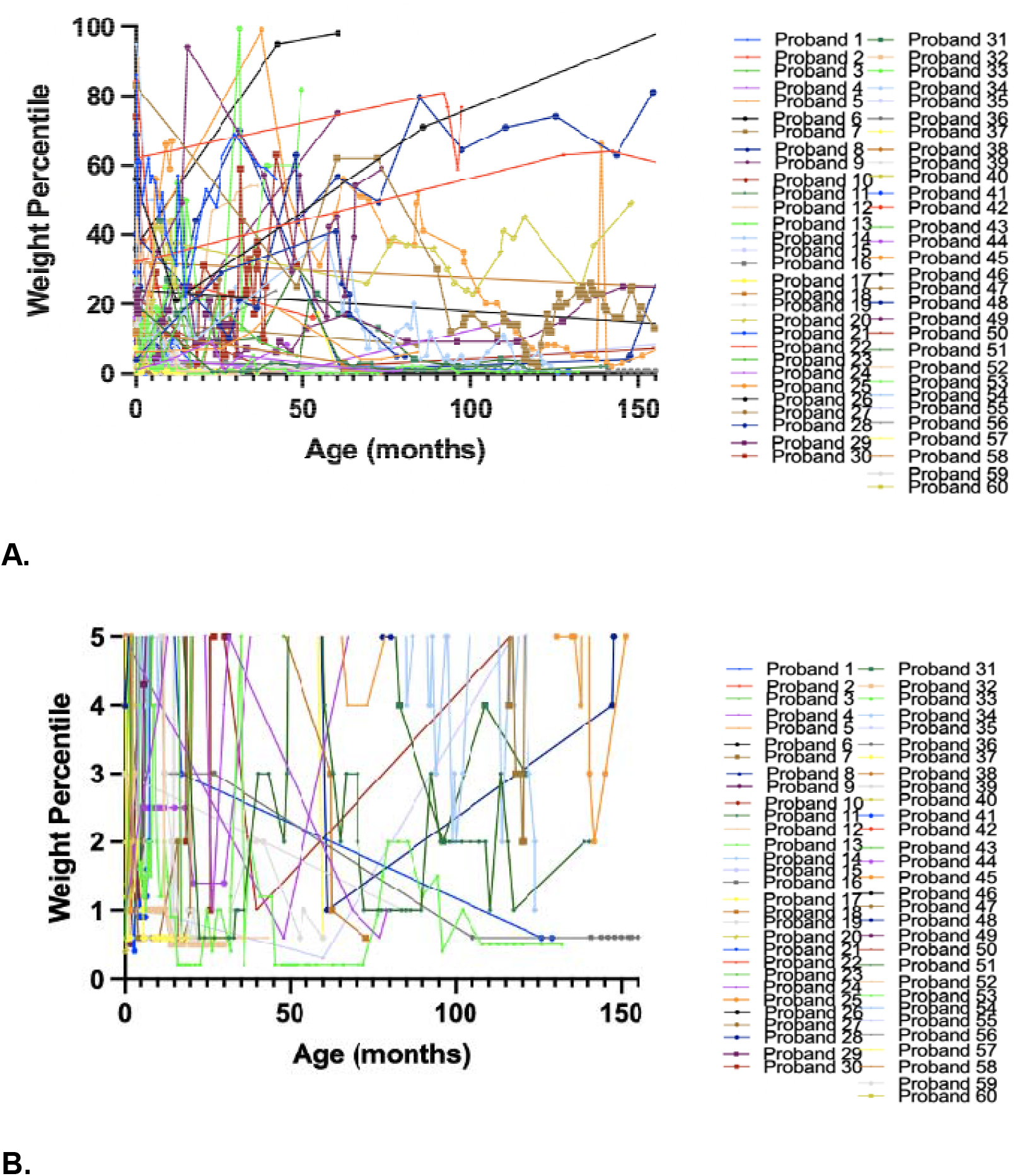
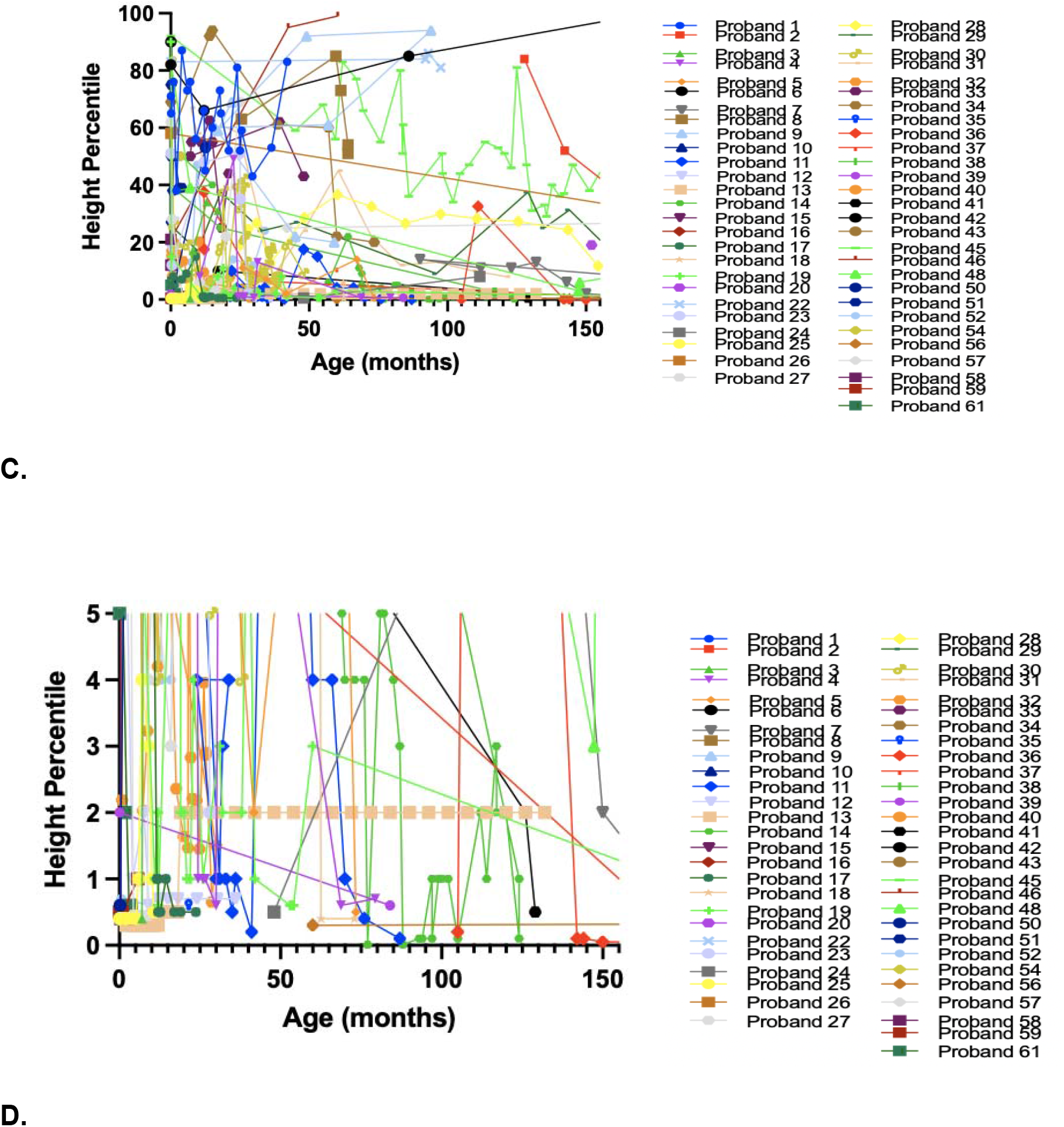
Weight and Height Trajectories of Ogden Syndrome Probands. **A**. Weight percentiles trajectories of Ogden Syndrome probands analyzed in this study (during the first 12 years of life). B. Depiction of probands whose weights were in percentiles characteristic of failure to thrive (below 3^rd^-5^th^ percentile) between birth and 12.5 years of age. C. Height percentile trajectories of Ogden Syndrome probands analyzed in this study (during the first 12 years of life). B. Depiction of probands whose heights were in percentiles characteristic of failure to thrive (below 3^rd^-5^th^ percentile) between birth and 12.5 years of age.

**Figure 2.**
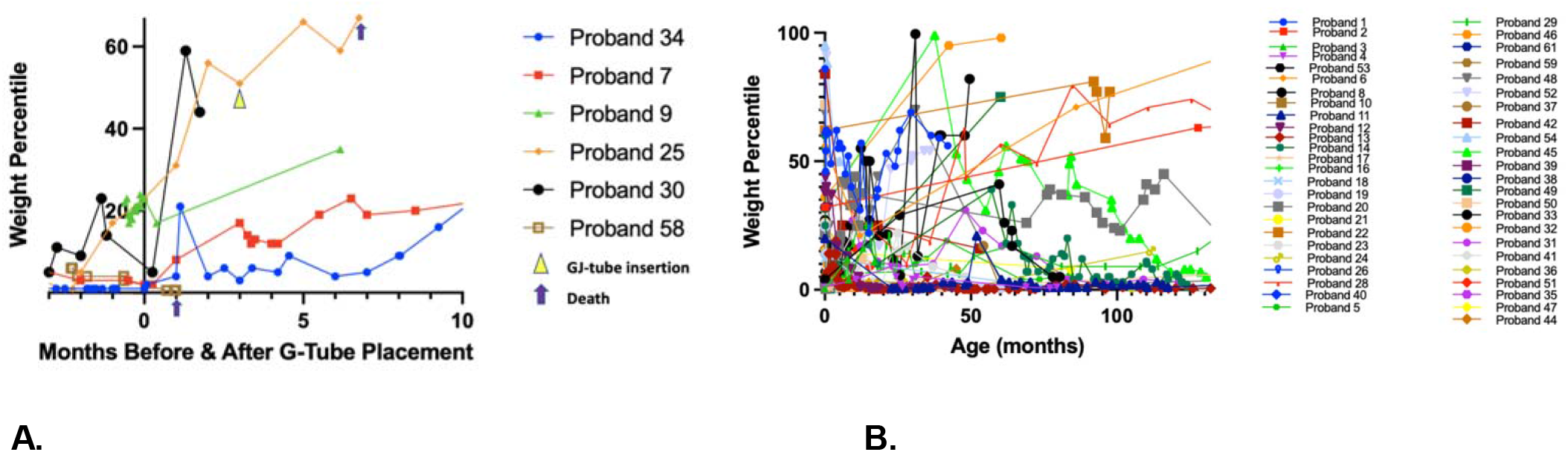
Gastrostomy Tube Efficacy. **A:** Weight percentiles of male and female probands before and after G-tube insertion. G-tube was inserted at x=0 for probands in this figure. All points to the left of zero on the x-axis correspond to weight percentiles prior to G-tube insertion, and all points to the right of zero on the -axis correspond to weight percentiles following G-tube insertion, while the G-tube was in place (and while utilized for nutrition by each respective proband). **Figure 2 B:** Weight percentiles of male and female probands who were never G-tube fed.

58% are at or below the 10^th^ percentile and 47% are at or below the 5^th^ percentile for weight (Figure 1C). 76% of data points between 0 and 30 months of age fall below the 20^th^ percentile. Also, of note, 72% of probands included in our study had at least one weight percentile value in the FTT range. Such weight percentile values confirm the previous findings of low weights among children with Ogden Syndrome. Similarly, although height percentile values demonstrate variability, 61% of data points are at or below the 10^th^ percentile for height and 29% of data points are at or below the 5^th^ percentile for height, indicating that the majority of analyzed probands are of small stature as compared to healthy age-matched children.

While variability of height and weight percentiles exists between probands, a substantial amount of variability is also seen on an individual basis. Healthy children typically remain in the same height and weight percentile throughout the majority of childhood. WHO growth charts are divided into the 0.1, 3^rd^, 15^th^, 50^th^, 85^th^, 97^th^, and 99.9^th^ percentiles. Although increases or decreases in height and weight percentile around 2-3 years of age may still indicate normal growth, before the age of 3, a height or weight percentile change that crosses two divisions (from the 3^rd^ to 50^th^ weight percentile for example) is deemed abnormal. After 3 years of age, healthy children should not experience growth percentile differences on a monthly or yearly basis until puberty (Marchand and Canadian Paediatric Society, Nutrition and Gastroenterology Committee 2012). However, our probands demonstrated dramatic height and weight percentile fluctuations during the first 3 years of life (Supplementary Figure 4A, C) and between 3 years of age and 11 years of age (Supplementary Figure 4B, D).

## Gastrointestinal Symptom Analysis

The following gastrointestinal symptoms were found in our probands: feeding difficulties in infancy (90%), dysphagia (75%), GERD (54%), vomiting (44%), constipation (34%), diarrhea/loose stools (24%), bowel incontinence (12%), eosinophils noted on endoscopy (10%) (Table 1). Supplementary Table 2 shows which symptoms each individual proband demonstrated.

These symptoms varied in severity, frequency, and nature. Feeding difficulties in early infancy included poor suck, poor swallow, no suck breath coordination, and frequent choking and gagging on breast milk or formula. Constipation ranged from intermittent to chronic and mild-moderate (relieved by Senna, lactulose, glycerin suppositories, or increased water intake) to severe (fecal impaction requiring ICU hospitalization). Several probands were reported to have occasional or recurrent loose stools without obvious diarrhea. Among children with diarrhea, frequency ranged from occasional bouts to frequent and chronic (6-7 foul smelling loose stools daily) and at times required hospitalization to treat dehydration or underlying gastroenteritis infection. Per parental reports, probiotics and reducing dietary sugar/fat content alleviated diarrhea in two of our probands.

For several children, GERD and silent reflux symptoms were described as constant, severe, or significant, while others had silent, mild, or moderate symptoms relieved by medications such as ranitidine and omeprazole. Barium swallow studies were not performed on all probands with GERD symptoms (or medical records for this imaging modality were unavailable) except for the following data: for Proband 17 with severe GERD, endoscopy and radiologic barium swallow reported that her sphincter muscle was underdeveloped, had low tone, and was more open at rest. Barium swallow imaging for Proband 26 showed “reflux from stomach into gullet. “ In our probands, dysphagia symptoms associated with difficulty swallowing foods (especially meat and bread) or fluids included: pain, coughing, gagging, choking requiring patting on the back or the Heimlich maneuver, and aspiration. While some of these probands had unremarkable X-ray barium sulfate swallow studies (or no study was performed), the three probands with abnormal findings all had oropharyngeal phase dysphagia. Of note, three probands with dysphagia also had ankyloglossia.

Vomiting varied among our probands, with different descriptors used, including severe, excessive, episodic, frequent (1-2x monthly), and daily (small emesis after dinner). For some probands, vomiting occurred without apparent reason, but in proband 7, this symptom was associated with menses and migraines and in proband 40 with low blood sugar and higher fat diet. One peculiar vomiting-related finding was noted in Proband 31; on an episodic basis, this female experienced 36-hour-long cycles of lethargy for 15-30 minutes, then 5 minutes of vomiting (filling “a couple of plastic bags full of food then mucus “ as per her parents), followed by a 15-30-minute asymptomatic period, after which the cycle repeated. Proband 38 was diagnosed with cyclical vomiting syndrome (CVS); beginning between 11 and 15 years of age, she vomited once monthly, which provoked a semi-comatose state according to her parents. The vomiting was associated with her menstrual cycle and triggered by indigestion-induced food build-up, tiredness, and certain foods such as pasta. Between 11 and 15 years old, Proband 48 was also diagnosed with CVS as well as abdominal migraine with vomiting at least twice a month. Symptom frequency reduced to once monthly following nortriptyline treatment. Triggers included “excitement, late nights, overeating restaurant food, and low-pressure weather events. “ Two probands with cyclic vomiting were reported to have Mallory Weiss tears, Probands 16 and 20 (3-4 times, leading to hematemesis). Of note, Proband 26, who had constant regurgitation (up to 3 hours after ingestion), experienced one episode of hematemesis when she was between 6 and 10 years of age, which required hospitalization.

Less commonly reported gastrointestinal symptoms included: bowel incontinence, esophageal or gastric dysmotility, delayed gastric emptying, bloating, recurrent abdominal distension (gaseous), celiac disease, cyclical bacterial overgrowths of small intestine, Giardia and C. diff infections, abdominal pain, celiac artery stenosis, dilated esophagus, severe subglottic stenosis, posterior glottic granulation tissue, vascular ring wrapped around esophagus, suspected abscess in caudate lobe of liver, and eosinophilic esophagitis. Only Proband 9 was formally diagnosed with Eosinophilic Esophagitis (EOE); however, four other probands had questionable EOE status due to presence of eosinophils on esophageal endoscopy. Endoscopy and esophageal biopsy results from one such proband (Proband 23), showed “squamous mucosa with moderate reactive changes, mild spongiosis, increased intraepithelial eosinophils (focally up to 12 eosinophils per high-power field). The reports determined that this degree of eosinophilia is somewhat borderline, and the differential diagnosis includes reflux esophagitis versus eosinophilic esophagitis. “ A sixth proband (Proband 60) demonstrated eosinophilic fibers of esophageal muscularis propria on autopsy.

Of the 61 children analyzed in our study, 9 children were G-tube fed at some point in life: four females and five males. All G-tube fed female probands demonstrated benefit from G-tube use as evidenced by parental testimonial or by increases in weight percentiles after tube insertion. Of note, four male G/GJ-tube-fed probands had died - Probands 25 and 28 (before 12 months of age) and Probands 30 and 60 (between 1 and 5 years of age).

Between 6 and 10 years old, Proband 7 began experiencing a steady decline in weight and started falling into a range diagnostic of failure to thrive (FTT), which is below the 3^rd^ to 5^th^ percentile (Figure 1A). Five days prior to G-tube insertion, she was in the 2^nd^ percentile for weight, but rose to the 8^th^, 17^th^, and 23^rd^ weight percentiles following roughly one month, two months, and six months, respectively status post G-tube placement and continued to track at or above the 20^th^ percentile for weight for over 18 months afterward. Her parent stated that the G-tube dramatically improved feeding for the patient, delivery of medications and hydration, as well as caregiving for the parents. They reported that “anybody with total care needs should get a G-tube, as it is a simple surgery, “ and that, “we truly wished we had placed the G-tube much earlier-if we knew then what we know now… “ Proband 7 ‘s exact feeding regimen is 250 calories daily from overnight supplemental feeds of Peptamen formula and 1000 calories from oral feedings. The child ‘s parent claims that “the extra 250 calories help. “ He/she also states that “even with the G-tube, you should not stop all oral feeds [as it is] better to give [children] some pleasure with oral feeds. “ For three months prior to G-tube insertion, Proband 34 was consistently falling in a weight percentile range diagnostic of failure to thrive (Figure 1 A). Less than one month following the beginning of G-tube use, the female ‘s weight percentiles rose to the point at which she no longer met criteria for FTT. Her weights kept increasing while being tube fed and within 10 months of tube feeding the patient rose from the 2^nd^ percentile for weight to the 24^th^. Similarly, the third tube-fed female proband analyzed demonstrated benefit; six months following G-tube insertion for Proband 9, she rose from the 21^st^ to the 35^th^ percentile for weight (Figure 1A).

For Proband 15, very few medical records were available to us, so we only have knowledge of her weight on one day between the ages of 11 and 15, but according to parental testimony in videoconference notes, this female “did gain weight after the tube, “ further supporting the efficacy of tube-feeding.

G-tubes were shown to be efficacious in four of the five male probands, whereas the fifth male proband demonstrated benefit from transitioning from a G tube to a GJ tube (which extends into the jejunal segment of the small intestine). For Proband 30, a G-tube was inserted due to inadequate PO intake. Peptamen Jr. formula was run through the tube. Approximately one month prior to placement, he was in the 14^th^ percentile for weight; about 1.3 months status-post onset of G-tube use, he reached the 59^th^ percentile for weight (Figure 1A). Similarly, Proband 27 experienced dramatic improvements while being G-tube fed with Peptamen Jr. This proband ‘s physician inserted a G-tube due to persistent vomiting of milk and mucus. Initially, Infatrin-Peptisorb was run through the tube. Prior to use of this formula, the proband showed an overall downward trend in standard deviations from mean weight of males his age, reaching 5.6 standard deviations below the mean 3.5 months before G-tube placement (Supplementary Figure 1A). At 2 months and 7 months of being fed Infatrin-Peptisorb through the G-tube, the proband ‘s weight became better (3.5 standard deviations below the mean). Although there was an improvement in terms of standard deviations, while on this formula, Proband 27 continued to track below the 1^st^ percentile for weight, and experienced dizziness and vomiting as per his parent. His formula was later changed to Peptamen Junior and within the same month of this change, the proband rose from 3.1 standard deviations below the mean to 2 deviations below the mean. While on this new formula, his weight trajectory had an upward trend and never reached values observed prior to G-tube placement. 9 months after starting the Peptamen Junior formula, the patient was at the 27^th^ percentile for weight, a striking improvement from being below the 1^st^ percentile for weight weeks prior to initiating this new formula. As per the proband ‘s parent, “the nutritionist recommended us Nestle Peptamen Junior…with this milk [he] is growing, gaining weight, he does not feel dizzy and is stronger. We are finally happy. “ They also stated, “with Peptamen Junior milk (1,5 Kcal/ml) [Proband 27] has notably improved his weight and height. “

Two male probands in our study were fed using a G-tube followed by a GJ-tube. Whereas Proband 25 experienced growth benefits from both, Proband 60 only showed improvements with the latter. On the day of G-tube insertion, Proband 25 was at the 23^rd^ percentile for weight, and he rose to the 56^th^ weight percentile during 2 months of G-tube utilization. Although medical records do not explicitly state this proband ‘s reason for transitioning from a G-tube to a GJ-tube, it was most likely due to persistent respiratory problems caused by reflux. Following GJ-tube placement and use of Monogen formula, the proband ‘s weight percentile trajectory demonstrated a continued upward trend (Figure 1A), reaching the 67^th^ weight percentile under 4 months status post GJ-tube insertion.

The second GJ-tube fed proband, Proband 60 experienced an overall increase in raw weight values following G-tube insertion (with Nissen fundoplication at the same operation, unlike any of the other probands).

However, he remained below the 1^st^ weight percentile while using a G-tube (Similac Advance formula) for 7.75 months. Physicians suspected that he was not adequately gaining weight due to his catabolic state from increased work of breathing; this proband was repeatedly admitted to the hospital for hypoxia and respiratory distress; he also had pectus excavatum deformity which was exacerbated during respiration, two small VSDs, and severe congenital scoliosis. His formula was switched to Nutren Jr. and caloric intake was increased, which reduced his number of standard deviations away from mean weight for boys his age, but kept this proband below the 1^st^ percentile for weight. 7.75 months after G-tube insertion, the tube was converted to a GJ-tube. Within 5 weeks of GJ-tube conversion, the child no longer met diagnostic criteria for FTT (Supplementary Figure 2). Whereas average monthly weight gain for a healthy child after 1 year of age is 8 oz, at a time between 12 and 16 months old, Proband 60 gained 38.8 oz in the one month following GJ-tube insertion. Prior to this, while with G-tube only, this child was gaining less than the expected number of ounces gained weekly by the average healthy child. It is also of note that prior to G-tube insertion, Proband 60 demonstrated a similar dramatic rise in weight after undergoing NG-tube insertion followed by NJ-tube transition within two weeks (Supplementary Figure 2A).

Proband 58 is the only child whose weight declined following G-tube insertion (Figure 1A). However, according to medical records, “He has had his G-tube two or three weeks. [Parent] thinks he is doing better than what he was before. “ Unfortunately, this proband died one month after G-tube placement. There is not enough data to conclude that the G-tube was ineffective for this proband with respect to weight percentiles. For Proband 60 and Proband 27, benefits from G-tube took over one month to demonstrate effect. Perhaps if Proband 58 had lived longer, the G-tube would have proved to be more efficacious.

For the children in this study who had mutations in the NAA10 gene but did not receive a G-tube or GJ-tube, there is no way of knowing whether their weight trajectories would have been more favorable had they been tube fed. However, whereas the majority of G or GJ tube fed probands had an upward trend in weight percentiles while having the tube in place (Figure 1A), there is tremendous variability in the weight percentiles corresponding to males and females who were not G or GJ tube fed with a sizeable portion of children localized in the FTT range (on or directly above the x-axis) (Figure 1B).

The efficacy of G-tubes and GJ-tubes with respect to growth poses a stark contrast to that of nasogastric (NG) tubes (Supplementary Figure 3A). Three male probands had NG-tubes only (their NG tube was not followed by a G or GJ tube). One of these children, Proband 35 remained below the 1^st^ percentile of weight for the 7.75 months while the NG-tube was in place. A second male in this category, Proband 59, had an NG tube inserted early in life (at an unknown date) and never reached above the 1^st^ percentile of weight in his lifetime (he died between 4 and 6 months of age). The final male in this group, Proband 56, experienced feeding difficulties in infancy. According to medical records, between birth and six months of age, he was well below [the] 3^rd^ percentile for weight for his age “ and “was not meeting his fluid and caloric intake requirements. “ At this time, he was diagnosed with varicella and had an NG-tube placed to supplement his feeds during his hospital stay. Although the varicella was uncomplicated and resolved, Proband 56 “fell considerably short of his requirements “ (he was at the 1^st^ percentile of weight) so his parents were advised to continue NG-tube feed supplementation at discharge. While using the NG-tube this child continued to demonstrate inadequate weight gain in his first 12 months of life, and when weighed at a visit between 3-5 years of age he was below the 0.4^th^ percentile for weight (Casey et al. 2015). Four out of the five male probands who were never NG-tube fed showed much higher weights as compared to boys who were NG-tube fed. Also, five male probands were NG-tube fed prior to G-tube insertion; none of these males rose above the FTT weight percentile range while the NG-tube was in place. Similarly, the weights of NG-tube fed girls tracked in low percentile ranges while having NG-tubes in place as well as months later (Supplementary Figure 3B).

## Discussion

Our weight data substantiates previous reports of children with Ogden Syndrome being small in terms of weight and stature, as evidenced by low percentile values. We also found that these values fluctuated dramatically on an individual level, often to an abnormal extent according to WHO criteria, suggesting that growth is disordered in multiple ways for this patient population. Additionally, we further confirmed that the phenotypic spectrum of Ogden Syndrome includes gastrointestinal pathology; almost every analyzed proband had at least one GI-related symptom. Just as the height and weight percentiles demonstrated variability, the GI symptom profile was heterogenous amongst our probands with respect to symptom frequency, severity, and nature. The clinical profile of OS was expanded to include findings that had not been previously described in the literature such as eosinophilic esophagitis, cyclic vomiting syndrome, Mallory Weiss tears, abdominal migraine, esophageal dilation, and subglottic stenosis. Previous studies have listed gastrointestinal findings in children with Ogden Syndrome, but there has been no mention of which symptoms were most prevalent. In our analysis, the five most common symptoms in order from most to least frequent were feeding difficulties in infancy, dysphagia, GERD/silent reflux, vomiting, and constipation. Although dysphagia among OS children has been reported in the literature, there has never been a specification of dysphagia type. We found that the three probands who had both dysphagia and abnormal findings on barium swallow imagining all had oropharyngeal dysphagia in particular.

Although our data is limited by the small quantity of probands analyzed, all boys and girls with NAA10 mutations displayed benefits from G-tube or GJ-tube feedings. For some of these children, G-tubes or GJ tubes were particularly helpful as a relatively quick method of raising weight percentiles out of the FTT range. For others who were tracking above the 3^rd^ to 5^th^ percentiles, G-tubes helped to increase weights to far healthier percentile ranges, at times crossing multiple growth chart divisions. Since the G-tubes or GJ-tubes were shown to be efficacious in some form for each child analyzed, recommendations for parents of children with Ogden Syndrome include G-tube insertion in case of low weight or severe feeding difficulty and not giving up on tube feeding if immediate, obvious growth benefits are not demonstrated. For two probands, although the G-tube was not showing clear growth improvements on one formula, after switching formulas (to Peptamen Jr. or Nutren Jr.) or increasing caloric intake (depending on caloric demands) they began to benefit dramatically in terms of growth values and/or parental testimonies. Caregivers should transition to a different formula or adjust caloric input through the tube and then closely monitor growth values before determining G-tube efficiency. If these two changes do not yield improvements in weights, parents should consider G to GJ tube exchange, a minimally invasive procedure that extends a catheter to the jejunum from the stoma that was already created for G-tube insertion (Kim et al. 2010), as the only two probands who underwent this exchange demonstrated substantial increases in weight following this conversion. With respect to children who did not show growth improvements from NG-tubes, although we cannot know how these children would have fared without these tubes (perhaps, theoretically they may have done worse without them), due to the growth benefits witnessed among children while being G-tube fed and the lack of improvements seen in weight among NG-tube fed children, another recommendation may be to forgo NG-tube placement and insert G-tubes during times of growth failure or severe feeding difficulties.

We also found that without G or GJ tube-feeding, probands with feeding difficulties and persistently low weights did not improve in weight as they became older. Despite often having undergone feeding therapy or having received adequate caloric intake, non-tube-fed probands who tracked in the FTT range for at least one year remained in this range for extended periods of time (often years), rarely ever reaching at least the 5^th^ weight percentile. For instance, Proband 11, who was never G-tube-fed and had trouble coordinating swallows and frequent choking, attended speech therapy twice weekly, incorporating mouth exercises to help with feeding. Also, her parents cut and blended her food, as well as added a thickener to make liquids easier to drink. Regardless, she weighed almost exclusively between the 0 and 3^rd^ weight percentiles for 9.8 years.

Similarly, in lieu of tube-feeding, Probands 12, 17, and 32 all worked with feeding therapists, who taught Proband 17 how to chew and strengthened Proband 17 ‘s feeding capabilities (as per her parent, who also supplemented this child ‘s diet with Similac Sensitive). However, after entering the FTT zone, they all stayed in the FTT range for at least 2.3 years. According to Proband 32 ‘s parent, the child ‘s “diet is still made up mostly of a real food formula, [including 800-900 calories from Kate Farms Pediatric standard formula daily] so we are able to track her calories fairly easily. By our calculations she is getting sufficient calories by mouth to facilitate adequate growth but obviously that is still not happening. “ Likewise, according to Proband 12 ‘s parents, she “get[s] many calories with 3 solid meals daily, “ also incorporating Kate Farms formula. For another OS child, Proband 13, “feeding had been a horrendous and painful battle pretty much her whole life, “ according to her parent who had considered G-GJ tube insertion, but opted to continue with oral feeds and “ridiculous amounts of calorie supplements through the day every day, but regardless she ‘ll be 13 years old [soon] yet she ‘s still in age 5 clothes, her older [sibling] without OS was in age 13 clothes by 9 years old and we ‘re fairly tall in our family… “ Since falling below the 5^th^ percentile for weight, Proband 13 has continued to have failure to thrive for 10.6 years. Although we have no data regarding the feeding therapies of Proband 36, we know that she had dysphagia, was never tube-fed, and had been in the FTT zone for ∼15 years. The aforementioned low-weight, non-tube fed probands with dysphagia were all female. Three of the lowest weighing male probands with dysphagia, no tube feeding, and no known feeding therapy were Probands 35, 59, and 60; the latter two boys died between birth and 12 months of age and spent their entire lives below the 5^th^ weight percentile. Proband 35 fell into the FTT range at 1 month old and continued to meet FTT criteria for ∼2 years. Please note that the previously mentioned durations in the FTT range (ex. 1.6 years (Proband 35), 15.3 years (Proband 36)) are all limited by our medical records, meaning Proband 35 did not grow out of the FTT range after 1.6 years, but rather his most recent available medical records end 1.6 years after he began displaying FTT weight percentiles.

A potential alternative to tube-feeding may be intensive efforts on the part of caregivers, as demonstrated by the case of Proband 3. This female tracked in the FTT range between birth and 5 months of life. Her speech language evaluation demonstrated poor range of motion and weakness of lips, tongue elevation of tip and in back, tongue lateralization, weakness of tongue retraction and widening, as well as poor mandibular range of motion. A barium swallow study performed in infancy was indicative of oropharyngeal dysphagia and included the following findings: pharyngeal weakness, swallow incoordination, aspiration during swallow, deep pharyngeal penetration, and nasopharyngeal reflux. Feeding tube insertion was recommended between 4-6 months old. Her parents decided to continue with oral feeding instead, even though getting the child to consume 4 oz. of high calorie formula took between 60-90 minutes. She ‘d nap for an hour afterward before it was time to feed her again because the process was so time-consuming. According to her parents, “basically, our life between feeding her and her [sibling] was like there was no in-between, we ended up getting a night nanny because it was so impossible…it was every waking hour. “ In terms of diet, they tracked their daughter ‘s calories and routinely added extra formula powder, egg yolk, and avocado to increase caloric intake. Within less than a month of beginning this meticulous regimen, Proband 3 was no longer in the FTT range, and until the present day has continued tracking above the FTT zone, even having reached the 25th percentile for weight when she was between 12 and 16 months old. (Supplementary Figure 5A). At ∼1 year-1.5 years old, this child ‘s parental efforts began to be supplemented with feeding therapy, which involved tongue strengthening exercises such as placing small pieces of food into the proband ‘s mouth and having her push them to the weaker side. Ultimately, she learned to chew and swallow without difficulty. However, it is worth noting that, as seen with other probands, although feeding therapy helped her learn how to eat properly, it did not lead to weight improvements (Supplementary Figure 5A).

An overall analysis of our study ‘s probands demonstrates that G-tube or GJ-tube feeding are effective measures for inducing weight gain, unlike NG-tube feeding. According to the case of Proband 3, intensive parental intervention involving consistent feeding, calorie tracking, and dietary supplementation as an alternative to tube-feeding may successfully treat growth failure. If such early efforts are not sufficient to raise an OS child ‘s weight above the 5th percentile by 6-12 months of age, parents should be strongly encouraged to consider G or GJ tube placement because continued caregiver efforts and enrollment in feeding therapy after this age have not been shown to engender weights above the FTT zone. For many probands in our study, weight data in the first year of life was not available. However, for four probands (13, 17, 32, and 43) who were not G/GJ-tube fed, weight data in the first year of life demonstrated that if parental efforts were not enough to supersede the FTT zone by 6 months of age, then FTT persisted for months to years afterward

(Supplementary Figure 6). This finding was substantiated by anecdotal evidence from the parent of Proband 4, who had dysphagia and was never G or GJ-tube fed. Although weight data for this child between birth and 10 months of age is not available, according to her parent, “she was and is still very small for her age…she ‘s always been a few years behind in sizing “ suggesting that in the first 10 months of life she demonstrated poor growth. Our records include her weight percentile data between 10 and 79 months of age, where she remained predominantly in the FTT range (Supplementary Figure 7) despite persistent parental attempts to feed her in spite of her frequent choking, their trials with different formulas (regular, Nutramigen, Elecare), thickening of formula with Simply Thick packets, a purely thickened-liquid-food diet (purees, oatmeals, cereals), and transition to finger feeding with solid foods.

It is worth highlighting that one proband, Proband 60, was the only child in our study who was fed with an NG-tube shortly followed by an NJ-tube. Unlike most boys and girls who did not show growth improvements while being NG-tube fed, Proband 60 displayed a dramatic rise in weight after NG-NJ conversion (Neosure formula). Unfortunately, however, despite its suspected benefits with respect to weight gain, an NJ tube is not recommended after further exploration of Proband 60 ‘s experience while using it. His NJ-tube became repeatedly clogged and the proband was intolerant of this tube, which caused him to frequently gag and spit up “increased amounts of thin brownish liquid “ as per his parents. According to his physician, “etiology of NJ tube feeding intolerance is likely due to reflux and aspiration possibly due to retraction of NJ tube back into the stomach or esophagus. “ So, his NJ tube was replaced with a G-tube, which did not help this child grow until it was transitioned into a GJ tube. Since this proband demonstrated his most substantial growth increases while on NJ and GJ tubes as compared to only NG and G-tubes (Supplementary Figure 2A), perhaps in children with Ogden Syndrome, growth failure is partially a consequence of absorption issues. NG and G tubes only bypass the stomach, whereas NJ and GJ tubes enter the jejunum; this may be a topic of discussion and exploration in future studies including more than two probands with jejunal tube feeding.

Another subject for further research should be the root of growth failure in OS children. As evidenced by our data, poor growth cannot be attributed to inadequate caloric intake, inability to properly chew or swallow, growth hormone deficiency, or low appetite; past the age of 6-12 months, calorie tracking, caloric supplementation, and using feeding therapy to successfully teach children how to chew, swallow, and no longer choke on food did not induce adequate weight gain. Low appetite was also not an observable cause of poor growth, as multiple proband parents claimed that their children had great appetites, were constantly hungry, and that “desire to eat has never been a problem. “ Growth hormone deficiency was treated with growth hormone administration in two of our probands (11 and 13), but these efforts kept these probands in the FTT range, although they did lead to weight improvements. A provocative test given for Proband 11 yielded the following results: peak GH to clonidine was 9.3 at 120 minutes and peak GH to arginine was 6.8 at 210 minutes. Of note, using a peak cutoff value of 6.8 µg/L accurately identifies 96.3% of patients with GH deficiency (GHD) (Guzzetti et al. 2016), and a cut-off of 3.3 µg/L produces a sensitivity value of 100% and 93% specificity to rule out GH deficiency among prepubertal children in particular (Silva et al. 2003). Among children demonstrating normal growth, average response to clonidine stimulation yielded a GH value of 13.1±1.8 µg/L with a range between 3.8 and 86 (Guzzetti et al. 2016). Although Proband 11 did not exactly meet the above criteria for GHD, after testing, she was subsequently treated with Norditropin (0.05 mg/kg QHS SC) at 6 years of age. According to Supplementary Figure 8, this proband ‘s weight demonstrated improvement, but she remained below the 3rd percentile for weight for at least 5 years while receiving GH treatment. GH treatment was terminated after the age of 12 for unclear reasons. Her parent deemed growth hormone to be “worthwhile and beneficial “ and stated that “we believe that the GH contributed to [Proband 11] following the growth curves rather than showing flat growth…we would recommend [GH treatment] to anyone who is concerned that their Ogden Syndrome child is not keeping up with the 0-3% growth percentile curves. Proband 13 was also treated with growth hormone, for an 11-month period between the ages of 5 and 10. This treatment was preceded by an arginine stimulation test result of 4.26 mcg/L which was deemed “suboptimal and supportive of partial growth hormone deficiency “ according to medical records. As per the parent of Proband 13, “growth hormone had little impact on increasing [Proband 13 ‘s] growth rate despite her being deficient. “ As previously stated, this proband had tracked in the FTT range for the past ∼11 years. While being treated with GH, this female stayed in the FTT zone, but did demonstrate a ∼1% increase in weight percentiles (Supplementary Figure 9). Effect of GH administration on weight gain among GH deficient OS children with growth failure should be further investigated in subsequent studies.

A first draft of the manuscript was written and circulated to the parents, and we received the following four responses. The parent of proband 34 wrote, “I can ‘t believe what some of these parents have done versus getting a G-tube. I wouldn ‘t have made it through all of this if it wasn ‘t for the G-tube. “ The parents of proband 3 replied to the draft stating that they made an error while reporting how long it took to feed proband 3 in infancy. Although they initially said that feeding duration was 60-90 minutes to consume 4 oz. of formula, a more precise estimate would be 20-30 minutes because the former duration included time spent feeding proband 3 ‘s sibling, and “prepping, diaper changing, clothes hanging, burping process, getting [their children] back to sleep, and then bottle cleaning. “ Also, the parents were specifically advised by a feeding therapist to not spend over half of an hour at a time bottle-feeding their child so as to avoid burning calories. Despite this, they note that “it was most certainly very intense, often spending 30 minutes feeding her a 4 oz. bottle that she sometimes didn ‘t finish “. They also claimed that when Proband 3 ‘s sibling (who does not have OS) was the same age as Proband 3, they finished their bottle “in just a matter of minutes. “

The third response received came from parents who had the following questions:

a. Did G/GJ tube feeding resolve GI symptoms such as reflux, diarrhea, and constipation?
b. How did tube-feeding affect developmental milestones in our analyzed probands?
c. What difficulties did caregivers face with respect to G/GJ tube maintenance?
d. Were there any more details regarding types of formula used and what were the night-time feeding regimens of tube-fed probands?

It is important to note that, initially, this study began with the question: are G-tubes efficacious with respect to improving gastrointestinal symptomatology in children with Ogden Syndrome. All available medical records were mined in search of an answer to this query. However, no documents explicitly detailed the consequences of tube-feeding on GI disturbances, so the benefits/drawbacks of tube feeding and their effects on weight were investigated instead in order to determine G/GJ tube efficacy. The only details regarding G-tube impact on GI symptoms came from the parent of Proband 27 in response to reading the first draft of this manuscript. They stated that their child continues to be G-tube fed with Peptamen Junior Advance hypercaloric milk formula (1.5 Kcal/ml) and “his constipation problems have improved a lot; “ they also provided new weight data, indicating that he is currently in the 40th percentile for weight.

Lack of sufficient data limits our ability to answer all of the aforementioned family ‘s questions, which should be thoroughly explored in future studies. All available information pertaining to formula types and feeding regimens were included in our paper, but unfortunately, for some probands, this information was not found in any medical records in our possession. Preliminary analyses of our data also entailed searching for the impact of tube feeding on developmental milestones and difficulty/ease of maintaining a G-tube, but little to no information was found. Proband 32 ‘s parents voiced concerns about G-tube maintenance, fearing that since their child is so mobile, a tube would constantly snag onto household or outside items, potentially leading to frequent irritation and infections. This raised the question of whether G/GJ tube-feeding is a hindrance to caregiving for OS children who actively move regularly. However, both the parents of Proband 7 (who was wheel-chair bound and persistently immobile) and the parent of Proband 34 (who walks and has no mobility issues) praised G and GJ tubes for significantly improving caregiving and did not mention any difficulty caring for and maintaining feeding tubes.

## Data Availability

All data produced in the present study are available upon reasonable request to the authors.

## Acknowledgements

This work is supported by New York State Office for People with Developmental Disabilities (OPWDD) and NIH NIGMS R35-GM-133408.

## Supplementary Tables

**Supplementary Table 1. Proband Number, cDNA, and Protein Mutations (see Excel sheet)**

**Supplementary Table 2. Gastrointestinal Symptoms of Each Proband (see Excel sheet)**

## Supplementary Figures

**Supplementary Figure 1.**
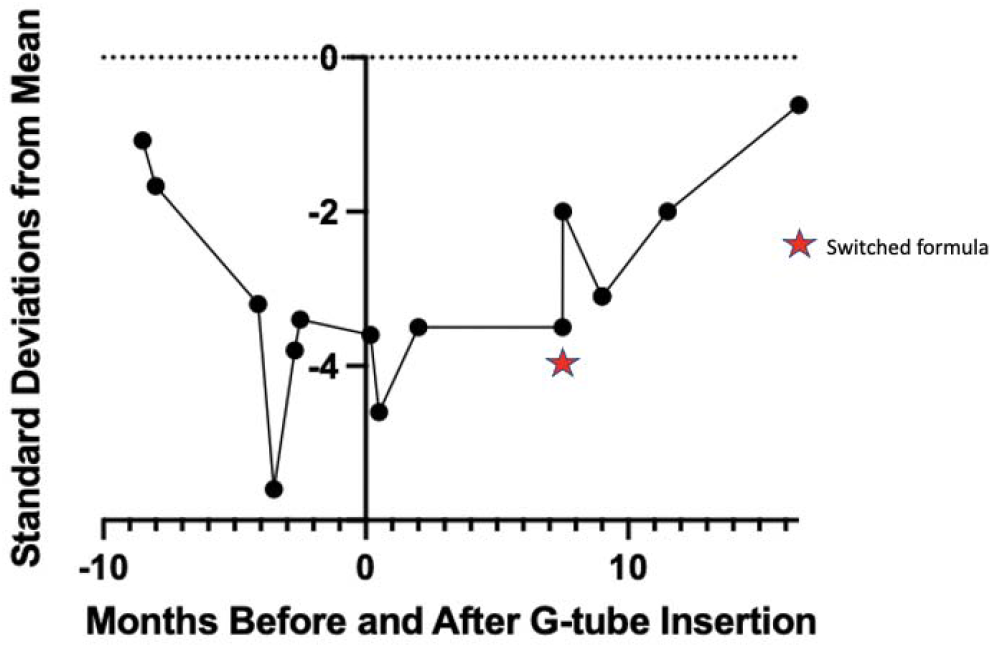
Weight trajectory of Proband 27 in terms of standard deviation from mean weight of healthy males his age.

**Supplementary Figure 2.**
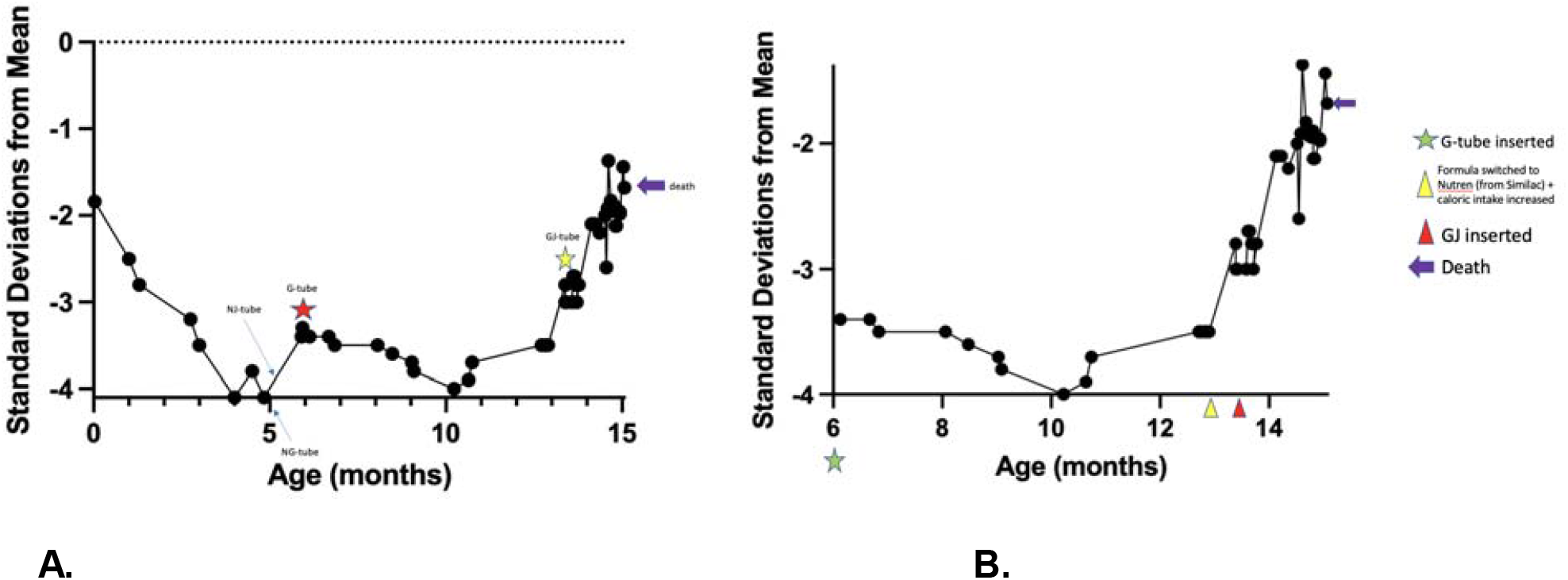
Proband 60 Weight Trajectories. A. Lifelong weight trajectory of Proba terms of standard deviation from mean weight of healthy males his age. B. Weight trajectory of 0 in Proband 60 in terms of standard deviation from mean weight of healthy males his age beginning after 6 months of age, at which point the proband had a G-tube inserted.

**Supplementary Figure 3.**
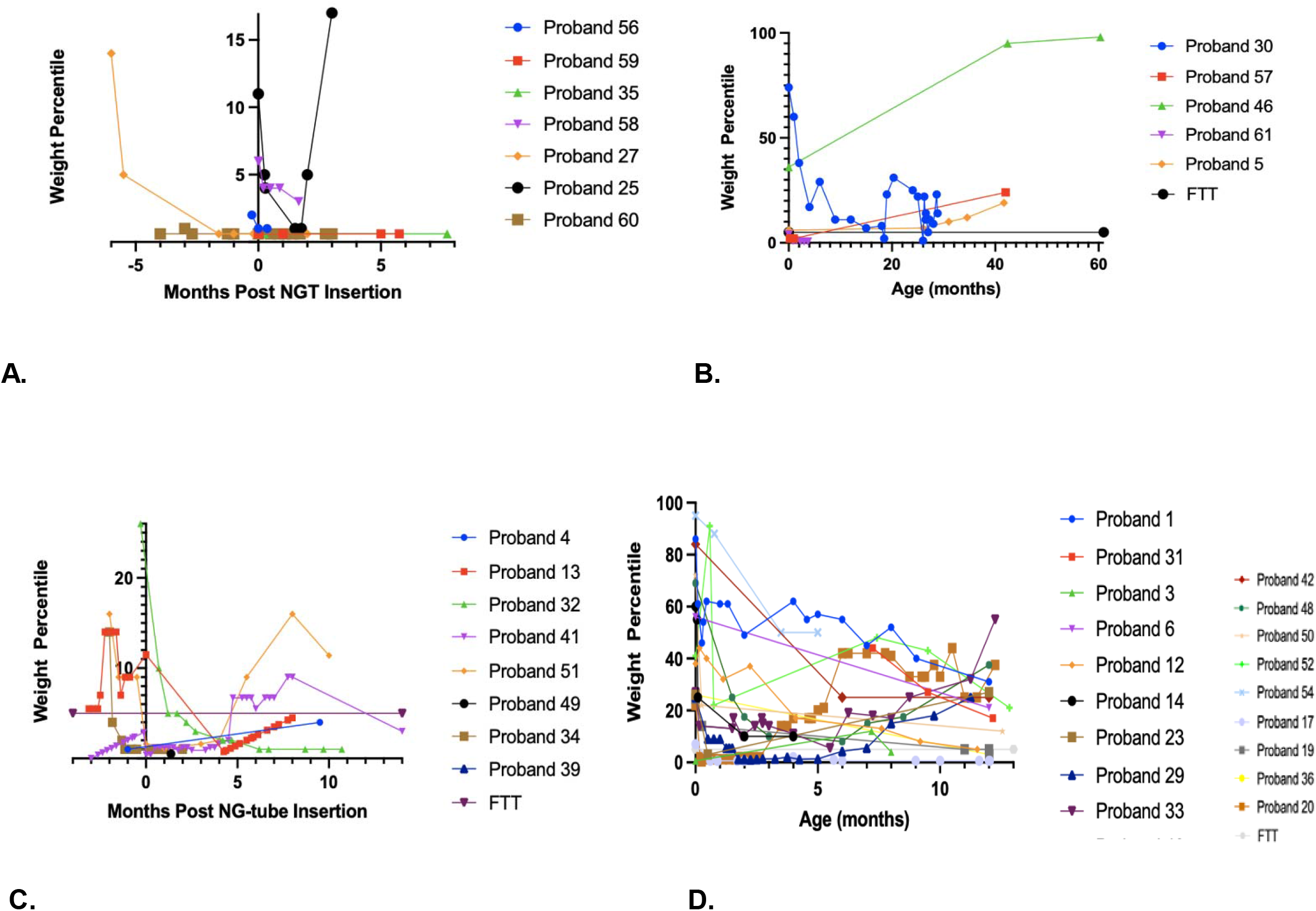
Weight trajectories of NG-tube-fed probands vs. probands who were never NG-tube fed. A. Weight trajectories of NG-tube fed male probands. B. Weight trajectories of male probands who were never NG-tube fed. C. Weight trajectories of NG-tube fed female probands. D. Weight trajectories of female probands who were never NG-tube fed.

**Supplementary Figure 4.**
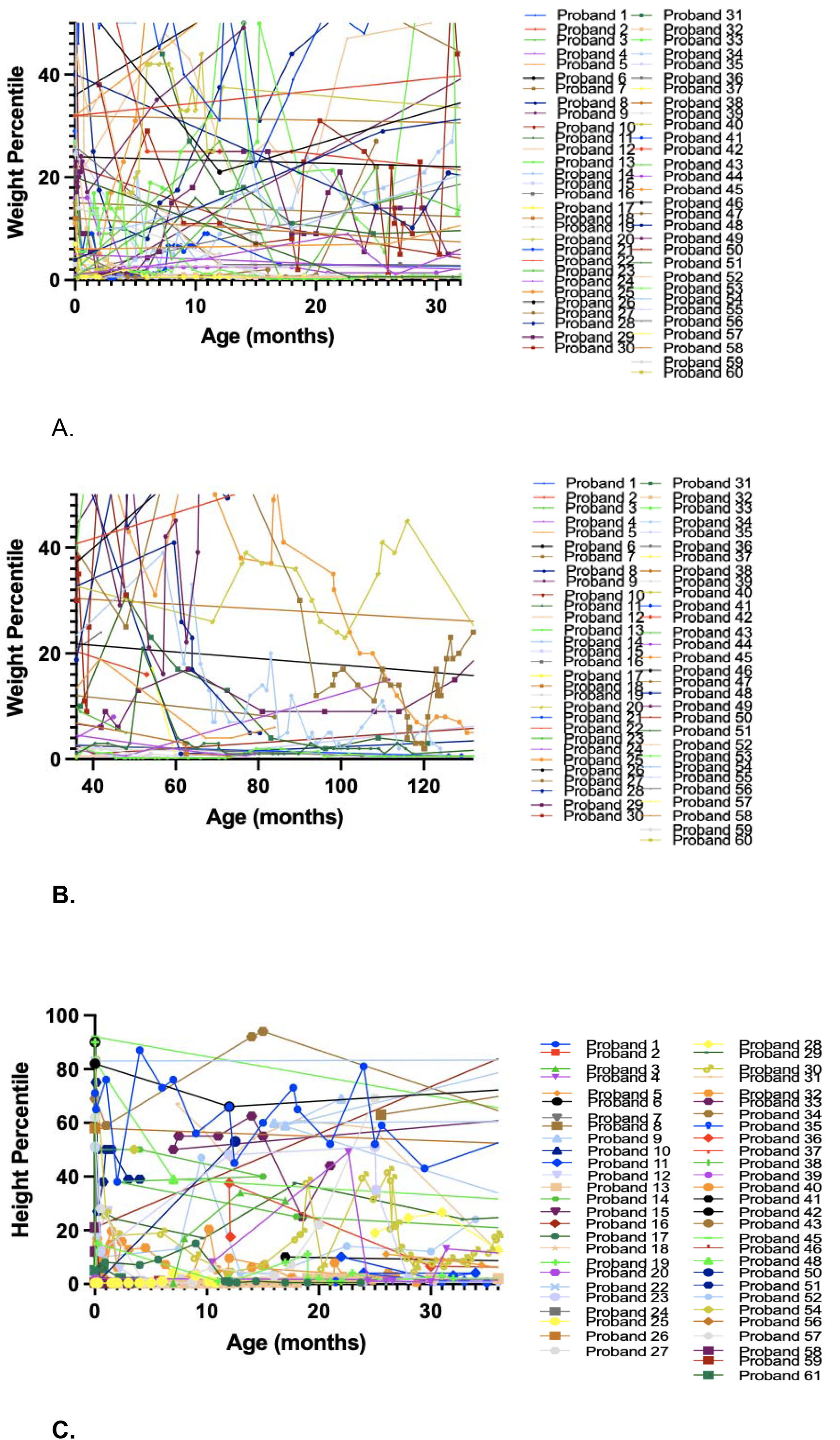
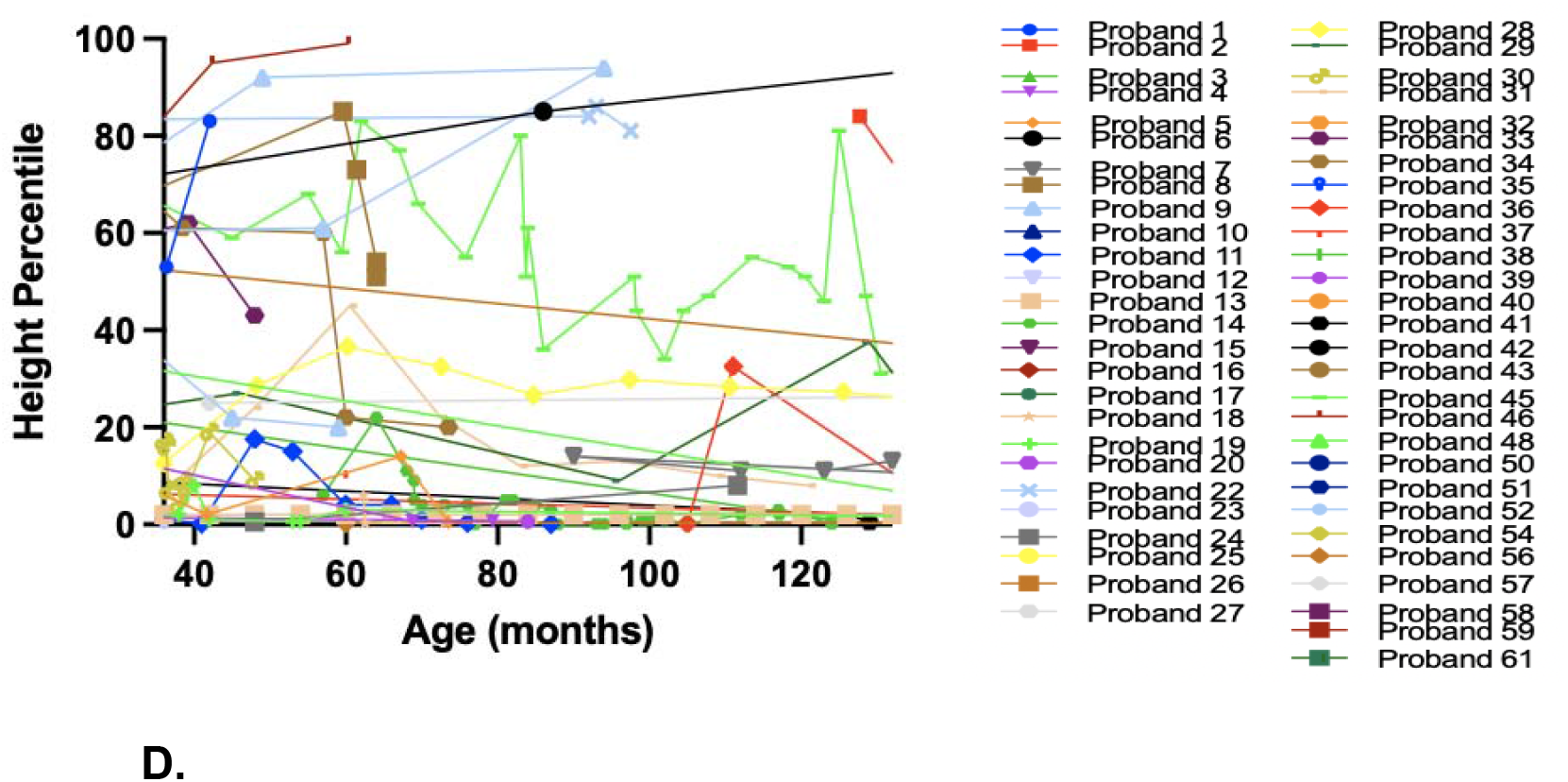
Height and weight percentiles at different age ranges. A. Weight percentiles during first 3 years of life. B. Weight percentiles between 3 and 11 years old. C. Height percentiles during first 3 years of life. D. Height percentiles between 3 and 11 years old.

**Supplementary Figure 5.**
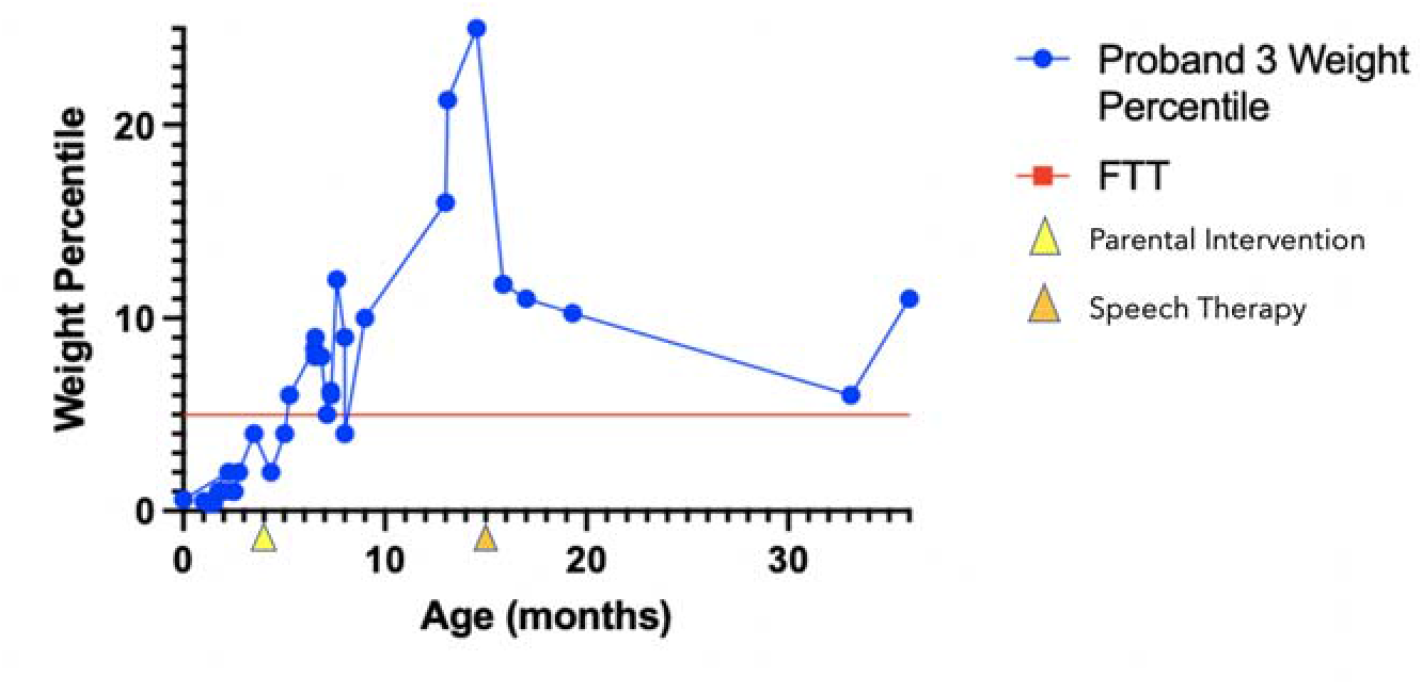
Weight Percentile Trajectory of Proband 3

**Supplementary Figure 6.**
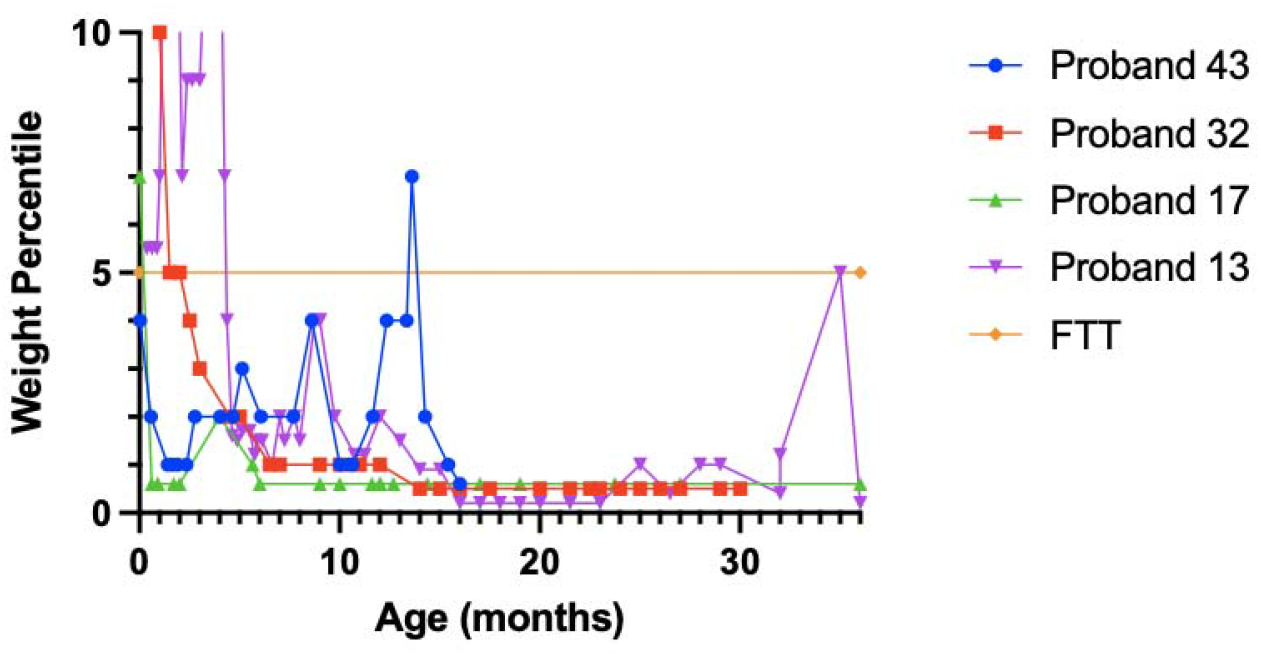
Weight Percentile Trajectories of Non-G/GJ-tube-fed Probands 13, 17, 32, and 43, in the first three years of life.

**Supplementary Figure 7.**
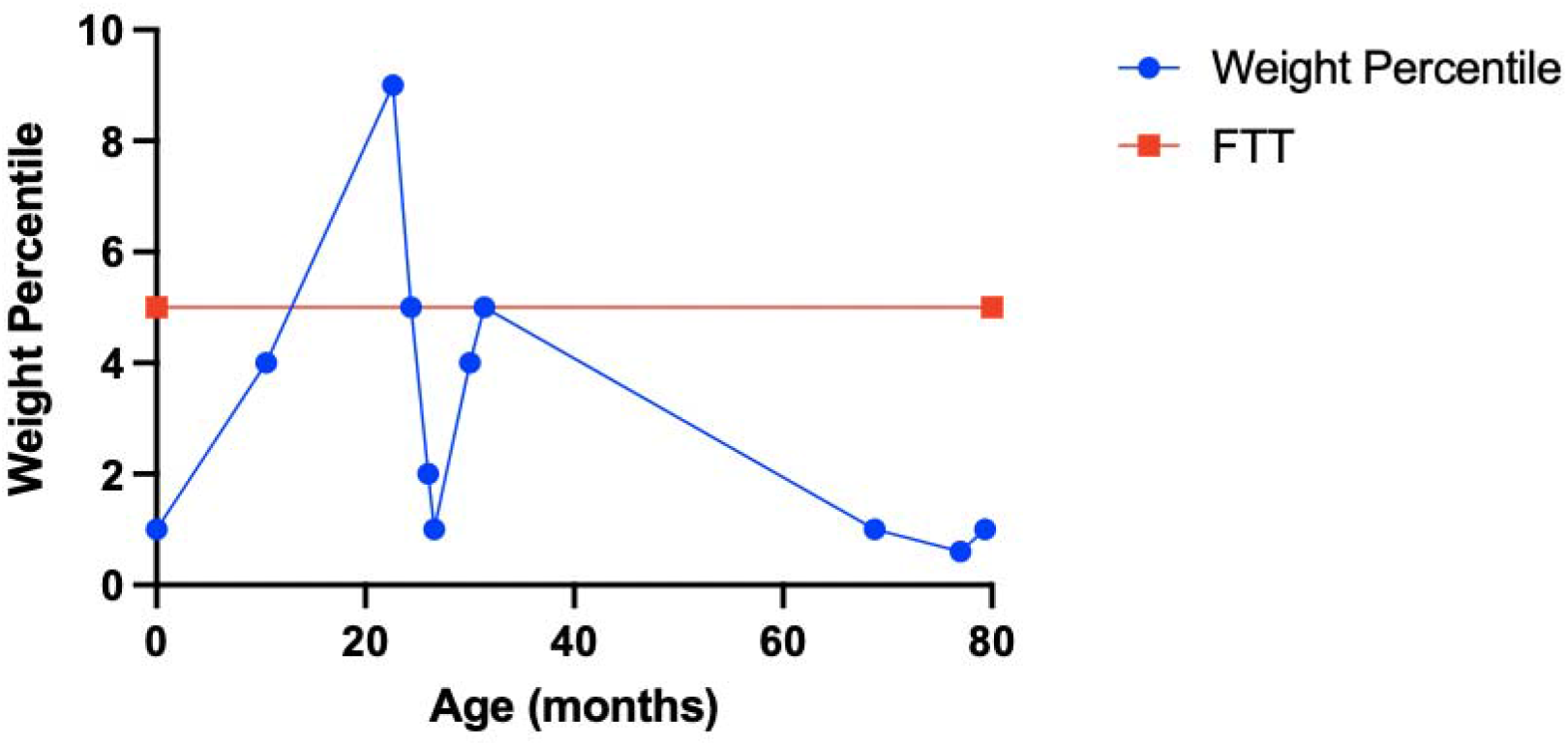
Weight Percentile Trajectory of Proband 4

**Supplementary Figure 8.**
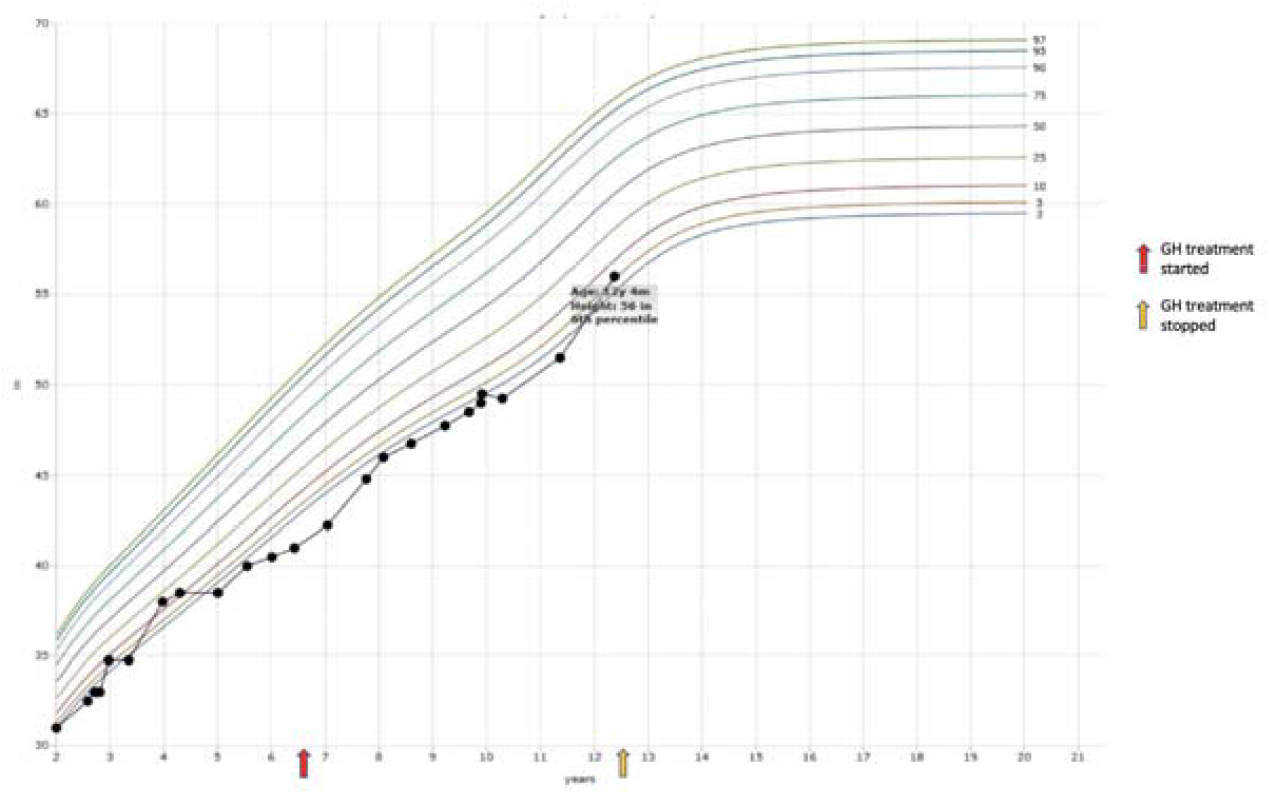
Growth Chart of Proband 11 including start and end of GH treatment

**Supplementary Figure 9.**
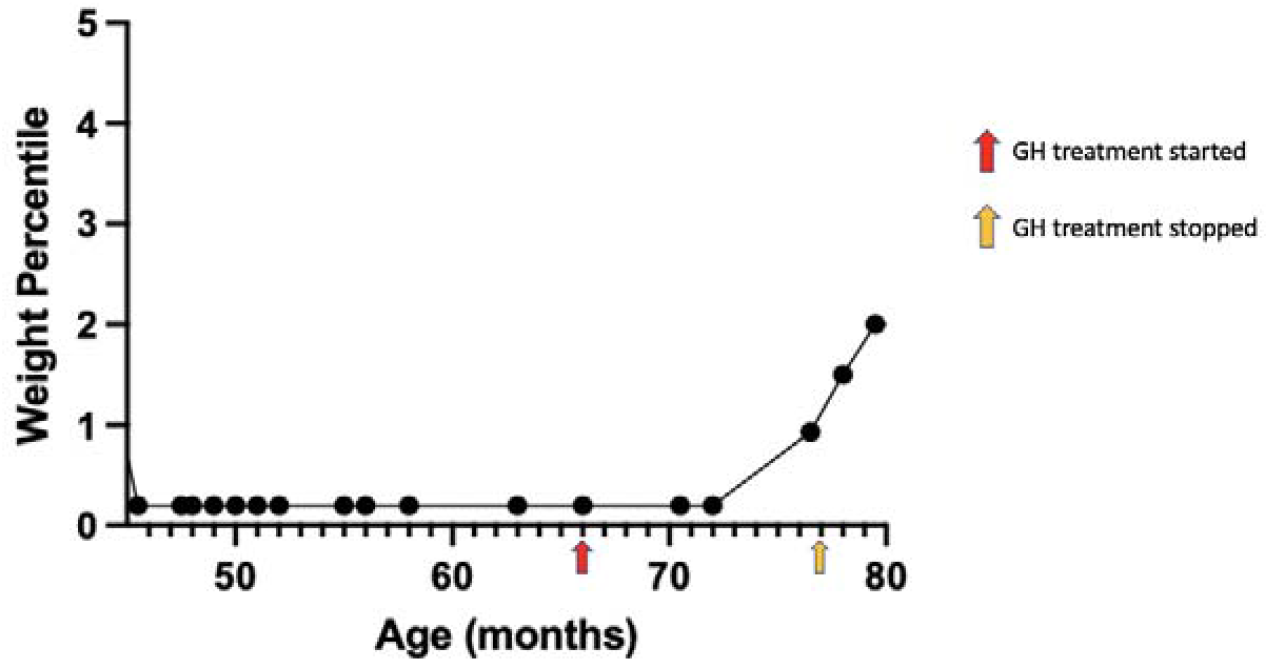
Weight Percentile Trajectory of Proband 13 before and after growth hormone Treatment

## Bibliography

Afrin, Antara, Jeremy W. Prokop, Adam Underwood, Katie L. Uhl, Elizabeth A. VanSickle, Roja Baruwal, Morgan Wajda, Surender Rajasekaran, and Caleb Bupp. 2020. “NAA10 Variant in 38-Week-Gestation Male Patient: A Case Study.” Cold Spring Harbor Molecular Case Studies 6 (6). https://doi.org/10.1101/mcs.a005868.

Bader, Ingrid, Nina McTiernan, Christine Darbakk, Eugen Boltshauser, Rasmus Ree, Sabine Ebner, Johannes A. Mayr, and Thomas Arnesen. 2020. “Severe Syndromic ID and Skewed X-Inactivation in a Girl with NAA10 Dysfunction and a Novel Heterozygous de Novo NAA10 p.(His16Pro) Variant - a Case Report.” BMC Medical Genetics 21 (1): 153.

Casey, Jillian P., Svein I. Støve, Catherine McGorrian, Joseph Galvin, Marina Blenski, Aimee Dunne, Sean Ennis, et al. 2015. “NAA10 Mutation Causing a Novel Intellectual Disability Syndrome with Long QT Due to N-Terminal Acetyltransferase Impairment.” Scientific Reports 5 (November): 16022.

Cheng, Hanyin, Leah Gottlieb, Elaine Marchi, Robert Kleyner, Puja Bhardwaj, Alan F. Rope, Sarah Rosenheck, et al. 2019. “Phenotypic and Biochemical Analysis of an International Cohort of Individuals with Variants in NAA10 and NAA15.” Hum Mol Genet. 2019 Sep 1;28(17):2900–2919. doi: 10.1093/hmg/ddz111.

Gogoll, Laura, Katharina Steindl, Pascal Joset, Markus Zweier, Alessandra Baumer, Christina Gerth-Kahlert, Boris Tutschek, and Anita Rauch. 2021. “Confirmation of Ogden Syndrome as an X-Linked Recessive Fatal Disorder Due to a Recurrent NAA10 Variant and Review of the Literature.” American Journal of Medical Genetics. Part A 185 (8): 2546–60.

Gupta, Angela S., Hind Al Saif, Jennifer M. Lent, and Natario L. Couser. 2019. “Ocular Manifestations of the NAA10-Related Syndrome.” Case Reports in Genetics 2019 (April): 8492965.

Guzzetti, Chiara, Anastasia Ibba, Sabrina Pilia, Nadia Beltrami, Natascia Di Iorgi, Alessandra Rollo, Nadia Fratangeli, et al. 2016. “Cut-off Limits of the Peak GH Response to Stimulation Tests for the Diagnosis of GH Deficiency in Children and Adolescents: Study in Patients with Organic GHD.” European Journal of Endocrinology 175 (1): 41–47.

Kim, Charles Y., Mayur B. Patel, Michael J. Miller Jr, Paul V. Suhocki, Anastasia Balius, and Tony P. Smith. 2010. “Gastrostomy-to-Gastrojejunostomy Tube Conversion: Impact of the Method of Original Gastrostomy Tube Placement.” Journal of Vascular and Interventional Radiology: JVIR 21 (7): 1031– 37.

Kweon, Hyae Yon, Mi-Ni Lee, Max Dorfel, Seungwoon Seo, Leah Gottlieb, Thomas PaPazyan, Nina McTiernan, et al. 2021. “Naa12 Compensates for Naa10 in Mice in the Amino-Terminal Acetylation Pathway.” ELife 10 (August). https://doi.org/10.7554/eLife.65952.

Lee, Chen-Cheng, Yi-Chun Shih, Ming-Lun Kang, Yi-Cheng Chang, Lee-Ming Chuang, Ramanan Devaraj, and Li-Jung Juan. 2019. “Naa10p Inhibits Beige Adipocyte-Mediated Thermogenesis through N-α-Acetylation of Pgc1α.” Molecular Cell 76 (3): 500-515.e8.

Maini, Ilenia, Stefano G. Caraffi, Francesca Peluso, Lara Valeri, Davide Nicoli, Steven Laurie, Chiara Baldo, Orsetta Zuffardi, and Livia Garavelli. 2021. “Clinical Manifestations in a Girl with NAA10-Related Syndrome and Genotype–Phenotype Correlation in Females.” Genes 12 (6): 900.

Marchand, Valérie, and Canadian Paediatric Society, Nutrition and Gastroenterology Committee. 2012. “The Toddler Who Is Falling off the Growth Chart.” Paediatrics & Child Health 17 (8): 447–54.

Mullen, J. R., P. S. Kayne, R. P. Moerschell, S. Tsunasawa, M. Gribskov, M. Colavito-Shepanski, M. Grunstein, F. Sherman, and R. Sternglanz. 1989. “Identification and Characterization of Genes and Mutants for an N-Terminal Acetyltransferase from Yeast.” The EMBO Journal 8 (7): 2067–75.

Polevoda, Bogdan, and Fred Sherman. 2003. “Composition and Function of the Eukaryotic N-Terminal Acetyltransferase Subunits.” Biochemical and Biophysical Research Communications 308 (1): 1–11.

Ree, R., L. M. Myklebust, P. Thiel, H. Foyn, K. E. Fladmark, and T. Arnesen. 2015. “The N-Terminal Acetyltransferase Naa10 Is Essential for Zebrafish Development.” Bioscience Reports 35 (5). https://doi.org/10.1042/BSR20150168.

Ree, Rasmus, Anni Sofie Geithus, Pernille Mathiesen Tørring, Kristina Pilekær Sørensen, Mads Damkjær, DDD study, Sally Ann Lynch, and Thomas Arnesen. 2019. “A Novel NAA10 p.(R83H) Variant with Impaired Acetyltransferase Activity Identified in Two Boys with ID and Microcephaly.” BMC Medical Genetics 20 (1): 101.

Rope, A. F., K. Wang, R. Evjenth, J. Xing, J. J. Johnston, J. J. Swensen, W. E. Johnson, et al. 2011. “Using VAAST to Identify an X-Linked Disorder Resulting in Lethality in Male Infants Due to N-Terminal Acetyltransferase Deficiency.” American Journal of Human Genetics 89 (1): 28–43.

Saunier, C., S. I. Stove, B. Popp, B. Gerard, M. Blenski, N. AhMew, C. de Bie, et al. 2016. “Expanding the Phenotype Associated with NAA10-Related N-Terminal Acetylation Deficiency.” Human Mutation 37 (8): 755–64.

Cheng, Hanyin, Leah Gottlieb, Elaine Marchi, Robert Kleyner, Puja Bhardwaj, Alan F. Rope, Sarah Rosenheck, et al. 2019. “Phenotypic and Biochemical Analysis of an International Cohort of Individuals with Variants in NAA10 and NAA15.” Human Molecular Genetics, May. https://doi.org/10.1093/hmg/ddz111.

Silva, Eveline G. P., Natasha Slhessarenko, Ivo J. P. Arnhold, Marcelo C. Batista, Vivian Estefan, Maria G. F. Osorio, Suemi Marui, and Berenice B. Mendonca. 2003. “GH Values after Clonidine Stimulation Measured by Immunofluorometric Assay in Normal Prepubertal Children and GH-Deficient Patients.” Hormone Research 59 (5): 229–33.

Støve, Svein Isungset, Marina Blenski, Asbjørg Stray-Pedersen, Klaas J. Wierenga, Shalini N. Jhangiani, Zeynep Coban Akdemir, David Crawford, et al. 2018. “A Novel NAA10 Variant with Impaired Acetyltransferase Activity Causes Developmental Delay, Intellectual Disability, and Hypertrophic Cardiomyopathy.” European Journal of Human Genetics: EJHG 26 (9): 1294–1305.

Wu, Y., and G. J. Lyon. 2018. “NAA10-Related Syndrome.” Experimental & Molecular Medicine 50 (7): 85.

